# Risk Assessments of COVID-19 Exposure of Government-owned Public Health Center Physicians According to WHO in Indonesia

**DOI:** 10.1101/2022.07.13.22277613

**Authors:** Anton Suryatma, Telly Purnamasari, Harimat Hendarwan

**Affiliations:** Research Centre for Pre-Clinical and Clinical Medicine, National Research and Innovation Agency of Indonesia

**Keywords:** risk, Public Health Center, physician, COVID-19, Indonesia

## Abstract

In 2020, the death of physicians due to COVID-19 in Indonesia raises questions about the condition that caused the incident. What was the situation at the early stage of the pandemic, the use of WHO’s Risk Assessment questionnaire, and what lesson was learned about it? A Cross-sectional survey, using blast mail surveys targeting the Government-owned Public Health Center Physician’s WhatsApp application across Indonesia had held. A Self-administered questionnaire, using WHO’s “Risk assessment and management of exposure of health care workers in the context of Covid-19” which has been translated into Bahasa Indonesia. As result, there were 2.099 responses eligible for this study. At the early stage of the pandemic, 99,29% of Government-owned Public Health Center Physicians were at high risk of COVID-19 exposure. Its because on average 64,23% of the respondent not use PPE correctly, 15,53% of respondents still performing actions that produced aerosols in health centers, or 22,73% of respondents got biological accidents. At the early stage of the pandemic in Indonesia marked by the scarcity of PPE, the lack of awareness from the physicians and or the government make a double burden on the physicians. As for the use of questionnaires, there were challenging issues in conducting the study, such as respondents feeling redundant in answering the questionnaire. It is recommended that the Central and Regional Governments, health centers, and hospitals increase their commitment to protecting physicians from possible exposure to Covid-19, among others, through meeting standard APD needs, maintaining the cleanliness of health care facilities, updating the skill and knowledge about pandemics on physicians, and providing adequate incentives. The physician is also expected to adapt in many ways.

## BACKGROUND

Indonesia reported the first case of Covid-19 on March 2, 2020. And declared a pandemic in April 2020. Until June-2020, Indonesia recorded 56.385 cases with 2,876 deaths. [1] [2]

Health workers who are at the forefront of handling COVID-19 cases have a high risk of contracting it. Health care workers have a substantial duty to diagnose and treat an exponentially growing number of patients. The World Health Organization (WHO) defines a health worker as all those involved in action whose primary purpose is to improve health, this includes physicians, nurses, midwives, paramedical staff, health care facility administrators, support staff, and community workers, who currently all face occupational risks of contracting COVID-19, and even death.

As of May 8, 2020, the number of deaths from health workers in Indonesia is in the top ten in the world. Indonesia ranks 7th with 55 health worker deaths, behind Ecuador (80 deaths), Iran (119 deaths), Russia (144 deaths), United Kingdom (163 deaths), United States of America (202 deaths), Italy (220 deaths). [3]

In Indonesia, until September 14, 2020, as reported by the Mitigation Team of the Indonesian Physicians Association (*Ikatan Dokter Indonesia*/IDI), there have been 115 physicians who died due to Covid-19, consisting of 7 professors, 57 general practitioners, and 51 specialists. [4]

The forefront of health facilities in Indonesia, in the majority, are Government-owned Public Health Centers (GPHC) also known as *Pusat Kesehatan Masyarakat* with the synonym of *Puskesmas*. In addition to GPHC, the provision of physician services is also in the practice of joint physicians, independent physicians, and hospitals.

As COVID-19 start in march 2020 in Indonesia, we conduct the survey in January 2021, we hope for the first 10 months of dealing with covid, we can assess COVID-19 exposure of GPHC Physicians using the WHO questionnaire. WHO launches interim guidance on risk assessment and management of exposure of health care workers in the context of COVID-19 in March 2020 with the purpose to help determine the risk of COVID-19 virus infection of all Health Care Workers who have been exposed to a COVID-19 patient and then provides recommendations for appropriate management of these HCWs, according to their infection risk. [5]

Ensuring the protection of health workers is important for every country as a strategic response to the COVID-19 crisis, especially when the government intends to understand more deeply the conditions at hand, as well as improve conditions and policies to minimize the incidence of illness and death of physicians due to Covid-19.

## METHODS

This study design is quantitative with a cross-sectional approach. The population in this study were all GPHC physicians in Indonesia, either their act as giving practice or at the managerial level. According to 2019 data, there are 10.203 GPHC throughout Indonesia, and approximately 24.750 physicians serve in GPHC. [6]

This study is in the form of a census therefore the sample of this study is the entire study population. Blast mail survey or survey by targeting a person’s online address was used, Questionnaire was expected to be filled out independently (self-administered questionnaire). The online address used is the WhatsApp application number from GPHC physicians across Indonesia. The delivery of messages starts from January 4 to January 6, 2021, then resumes from January 15 to 19, 2021. The filling time is set to end on January 25, 2021.

The WHO questionnaire “Risk assessment and management of exposure of health care workers in the context of COVID-19”, which was translated into Indonesian and added some questions, was used. The essence of the WHO questionnaire is to classify respondents into high and low-risk exposure to covid and variables are the use of PPE, handwashing behavior, PPE removal, and biological accidents. According to WHO, high risk is if the respondent does not answer ‘always’ on the question of the use of PPE and preventive behavior also and/or answers ‘yes’ on the question of biological accidents, while the low risk is other than those mentioned.

## RESULT

### The Challenges of the study

Indonesia is a very big country with more than 17.000 islands spread across the nation. Because we are using the WhatsApp application to send messages, and the application needs a mobile signal, and not all areas were covered by a mobile signal, then it became a challenge to reach out to the respondents. This challenge has been overcome by parallel instructing the Provincial Health Office and the District Health Office to reach the GPHC physician.

The next challenge is the respondents feel redundant in answering the question. No. 5 and 6 of the questionnaires are similar to each other. We overcome the problems by explaining to the respondent that the two number on the questionnaire is different and asking the respondent to read carefully.

**Figure 1.**
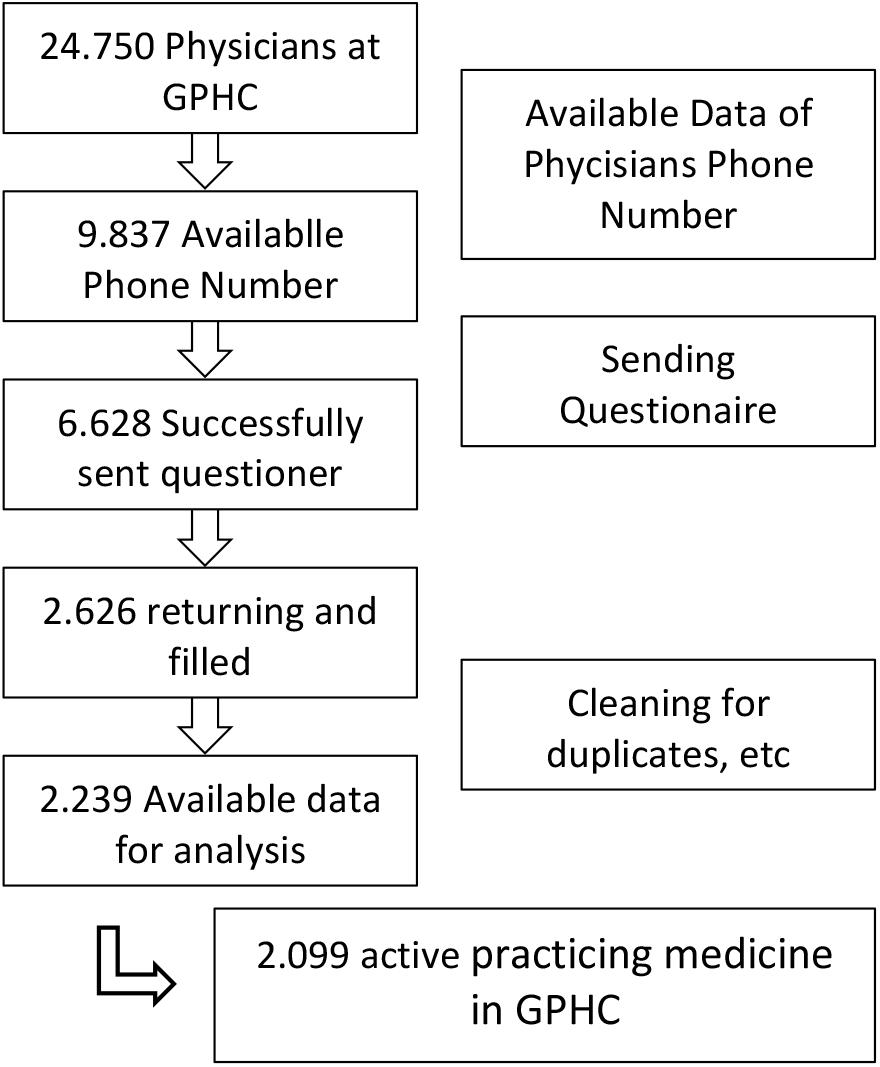
Flowchart of Respondent.

### Characteristic

A total of 6.628 Government-owned Public Health Center mobile phones were successfully contacted, and 2.626 numbers were returned to the questionnaire. After the cleaning process (multiple numbers are issued (only choose the one with the most complete filling), non-physician respondents, and respondents who do not include their title), there were 2.239 data processed. Respondents were spread across 34 provinces (all the provinces in Indonesia). The proportion of male respondents was 29,61% (663) and women 70,39% (1.576). The age range of respondents ranged from 23 years to 62 years with an average age of 35,5 years.

Of the total respondents, there were 45 people (2,01%) respondents who did not practice medicine in a government-owned public health center (GPHC). All of those who do not practice medicine are the head of the health center and medical services at the health center were done by other staff physicians.

Related to the activeness of physicians in handling COVID-19, it can be seen in table 1, almost entirely from physicians who practice also carrying out COVID-19 handling services. However, there are some physicians, 95 people (4,24%), who despite practicing medicine, are not involved in handling COVID-19. This is on average due to the local policy of the health center not to give responsibility for handling COVID-19 to the physician due to age or health conditions.

**Table 1.** HCW activities performed on COVID-19 patients in a health care facility

| No. | Question N0 4 | N | Yes (n/%) |
| --- | --- | --- | --- |
| A | Provide direct care to a confirmed COVID-19 patient? | 2.239 | 2.099 / 97,99 % |
| B | have face-to-face contact (within 1 meter) with a confirmed COVID-19 patient in a health care facility? | 2.099 | 1.534 / 73,08 % |
| C | Present when any aerosol-generating procedures were performed on the patient | 2.099 | 326 / 15,53 % |
|  | Types of procedure* |  |  |
|  | - Tracheal Intubation | 307 | 7 / 2,28 % |
|  | - Nebulizer treatment | 315 | 193 / 61,27 % |
|  | - Open airway suctioning | 306 | 36 / 11,76 % |
|  | - Collection of sputum | 306 | 108 / 35,29 % |
|  | - Tracheotomy | 308 | 0 / 0 % |
|  | - Bronchoscopy | 307 | 0 / 0 % |
|  | - Cardiopulmonary resuscitation (CPR) | 304 | 64 / 21,05 % |
| D | Has direct contact with the environment where the confirmed COVID-19 patient was cared for (eg: bed, linen, medical equipment, bathroom, etc.) | 1936 | 594 / 30,68 % |
| E | Involved in health care interaction(s) (paid or unpaid) in another health care facility during the period above | 2099 | 980 / 46,69 % |
|  | Added question about workplaces** |  |  |
|  | - Hospital | 445 | 152 / 34,16 % |
|  | - Clinic / Joint Practice | 557 | 332 / 59,61 % |
|  | - Private Practice | 769 | 651 / 84,66 % |

\*population of saying yes in performing aerosol-generating procedures and excluded missing data
\*\*questions added on the original version

There are physicians who in addition to working in health centers, then he practices elsewhere such as in hospitals (154 people), joint practices (335 people), or private practices (647 people).

### Activities performed on COVID-19 patients including aerosol-generating procedures

From the question about often face-to-face with COVID-19 patients (within 1 meter), more than half of respondents stated that they often do this (1.534 / 73,08%). And for the aerosol-generating procedures, most of the respondents stated that they never do aerosol-generating procedures in GPHC (1.773 people / 84,47%). Of the procedures performed, there were more procedures in the form of nebulizer (193 people / 16,3%), followed by sputum collection (108 people / 6.4%) and cardiopulmonary resuscitation (64 people / 21,05%).

### Infection prevention and control (IPC) during health care interactions

In the questionnaire, it was asked about 4 Personal Protective Equipment worn by physicians in Covid-19 services and whether to replace PPE according to the protocol. The complete results can be seen in table 2. One of the main health protocols is the use of masks, in this study the use of N95 masks in preventing Covid-19 transmission, only 33% of GPHC physicians. If we take the average proportion of the always using PPE in the respondent {(62,84+96,00+52,88+45,21)/4}, then only 64,23% of Physicians who always wear the PPE.

**Table 2.** Adherence to IPC procedures during health care interactions

| No. | Question No. 5 | 1* | 2* | 3* | 4* |
| --- | --- | --- | --- | --- | --- |
| A | Wear personal protective equipment (PPE) during a health care interaction with a COVID-19 patient |  |  |  |  |
|  | 1. Single-use gloves | 1319<br>(62,84) | 393<br>(18,72) | 275<br>(13,10) | 102<br>(4,86) |
|  | 2. Medical mask | 2015<br>(96,00) | 63<br>(3,00) | 11<br>(0,52) | 2<br>(0,10) |
|  | 3. Face shield or goggles/protective glasses | 1110<br>(52,88) | 498<br>(23,73) | 344<br>(16,39) | 137<br>(6,53) |
|  | 4. Disposable gown | 949<br>(45,21) | 432<br>(20,58) | 381<br>(18,15) | 303<br>(14,44) |
| B | Remove and replace PPE according to the protocol (eg: when medical mask became wet, disposed of the wet PPE in the waste bin, performed hand hygiene, etc.) *during a health care interaction with the COVID-19 patient* | 1595<br>(75,99) | 338<br>(16,10) | 101<br>(4,81) | 50<br>(2,38) |
| C | Perform hand hygiene before and after touching the COVID-19 patient regardless of wearing gloves *during a health care interaction with the COVID-19 patient* | 1871<br>(89,14) | 154<br>(7,34) | 13<br>(0,43) | 9<br>(0,43) |
| D | Perform hand hygiene before and after any clean or aseptic procedure (eg: while inserting a peripheral vascular catheter, urinary catheter, intubation, etc.) *during a health care interaction with the COVID-19 patient* | 1757<br>(83,71) | 141<br>(6,72) | 47<br>(2,24) | 46<br>(2,16) |
| E | Perform hand hygiene after exposure to body fluid *during a health care interaction with the COVID-19 patient* | 1917<br>(91,33) | 91<br>(4,34) | 25<br>(1,19) | 10<br>(0,48) |
| F | Perform hand hygiene after touching the patient's surroundings (bed, door handle, etc.), regardless of wearing gloves *during a health care interaction with the COVID-19 patient* | 1654<br>(78,80) | 295<br>(14,05) | 82<br>(3,91) | 13<br>(0,62) |
| G | Were high-touch surfaces decontaminated frequently (at least three times daily) *during a health care interaction with the COVID-19 patient* | 852<br>(40,59) | 634<br>(30,20) | 398<br>(18,96) | 160<br>(7,62) |

\*Presented as n (%) with 1=Always, as recommended; 2=Most of the time; 3. Occasionally; 4. Rarely

### Infection prevention and control (IPC) when performing aerosol-generating procedures

The questionnaire asked about 6 behaviors related to prevention during aerosol-generating procedures on a COVID-19 patient. namely wearing and removing PPE according to protocols, doing hand hygiene or washing hands before and after touching a Covid-19 patient, before and after performing aseptic procedures, after touching objects around the patient, and decontaminating the surfaces of objects touched.

The behavior of washing hands in the GPHC is classified as good. On average, 80% of respondents do this. The behavior that is not carried out according to this questionnaire is the decontamination of the surfaces of objects that are often touched with less than 50% who do it.

### Biological Accident

Most of the respondents stated that they had not experienced a biological accident at work, 77.3% had never experienced one at the GPHC and 81.4% had never experienced it at work other than at the GPHC (table 4). However, there are still 22.7% of physicians who experience it at the GPHC, and the most frequent form of biological accident is the splashing of secretions from the mouth/nose.

**Table. 3.**
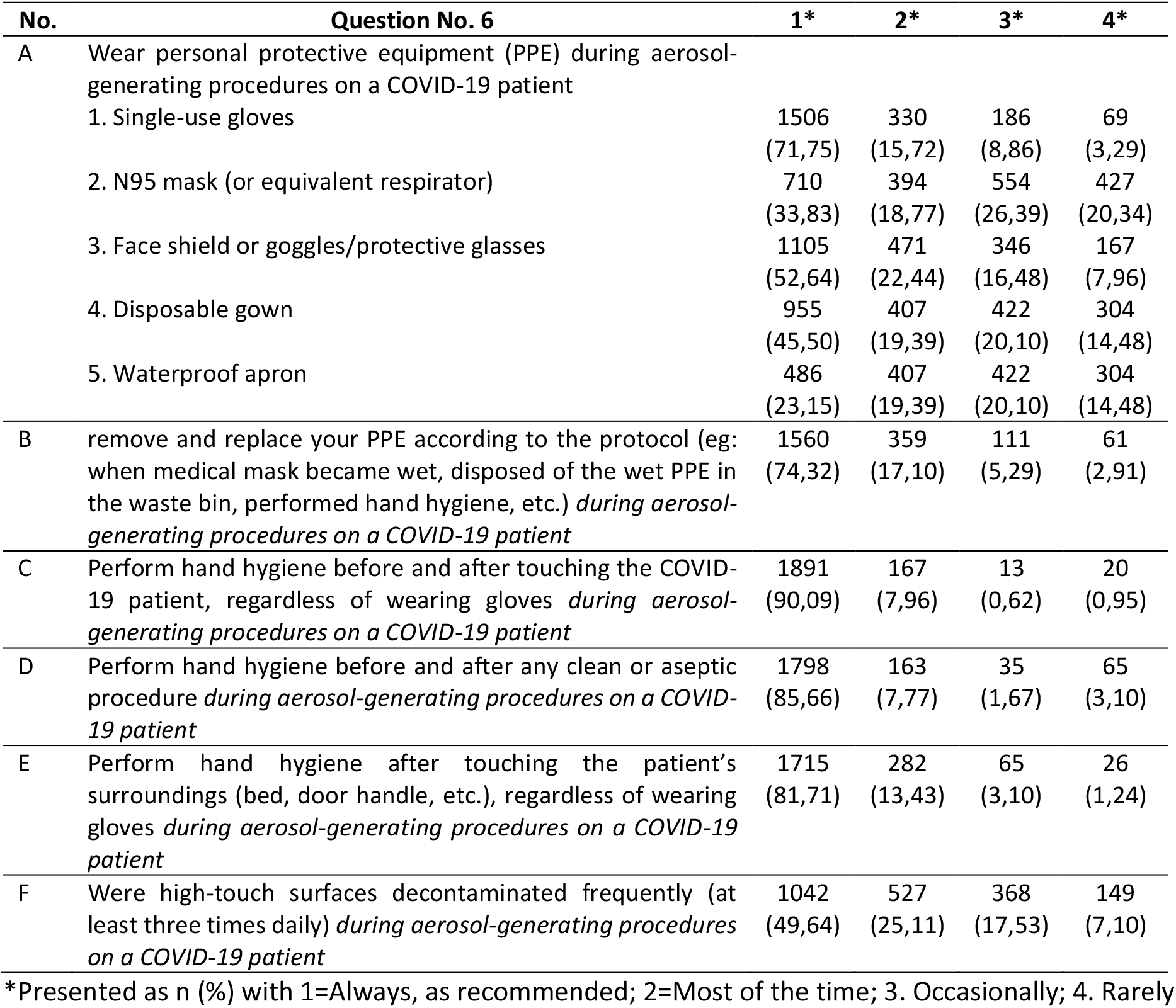
Adherence to IPC measures when performing aerosol-generating procedures

**Table. 4.** Biological Acccident

| No | Question No. 7 | N | Yes (n / %) |
| --- | --- | --- | --- |
|  | Types of accident |  |  |
|  | - Splash of biological fluid/respiratory secretions in the mucous membrane of eyes | 477 | 135 (28,30) |
|  | - Splash of biological fluid/respiratory secretions in the mucous membrane of mouth/nose | 477 | 257 (51,36) |
|  | - Splash of biological fluid/respiratory secretions on nonintact skin | 477 | 70 (14,68) |
|  | - Puncture/sharp accident with any material contaminated with biological fluid/respiratory secretions | 477 | 27 (5,66) |

**Table. 5.** WHO Risk Assessment Categorization

| Labels | High Risk | Low Risk |
| --- | --- | --- |
| **No. Risk (question No. 5)** | **16** | **15** |
| **No-Risk (question No. 6)** | **2** | **15** |
| No-Risk (question No. 7) |  | 15 |
| Risk (question No. 7) | 2 |  |
| **Risk (question No. 6)** | **14** |  |
| No-Risk (question No. 7) | 14 |  |
| **Risk (question No. 5)** | **2068** |  |
| **No-Risk (question No. 6)** | **77** |  |
| No-Risk (question No. 7) | 68 |  |
| Risk (question No. 7) | 9 |  |
| **Risk (question No. 6)** | **1991** |  |
| No-Risk (question No. 7) | 1882 |  |
| Risk (question No. 7) | 109 |  |
| **Grand Total** | **2084 (99,29%)** | **15 (0,71%)** |

### Risk Classification

According to the classification that accompanies this questionnaire, it can be seen that 99% of doctors in Indonesia are categorized as high risk.

Interestingly, two people are good at prevention and behavior but unfortunately got a biological accident, which makes them in the high-risk category.

## DISCUSSION

### The use of the WhatsApp application in Indonesia

Approximately in 2020, there are 272 million population in Indonesia with 338 million mobile phone connections and 160 million active social media users. The second social media most often accessed in Indonesia is WhatsApp, the first is YouTube, and it makes WhatsApp the first messaging application in Indonesia, defeating the conventional cellular provider messaging application. Users of this messaging application reached 134.4 million users in Indonesia [7]. With this number of users, WhatsApp is considered the best platform to reach out to Indonesian respondents. The problem is that sometimes respondents do not put or change their WhatsApp numbers in their data, so even though the penetration is high, it also needs an updated database about the respondent number.

### Indonesian physicians dan COVID-19

From the results of the characteristics of this survey, it is known that almost all GPHC physicians are involved in handling covid, this is also proven by the average number of doctors working in one GPHC is 1 person. [8]. This means that the burden on GPHC physicians will increase when the epidemic hits. In handling COVID-19, the role of physicians in diagnosing COVID-19 requires taking a patient history, in this case, direct contact with the patient. In this study, more than half of doctors made contact at a distance of less than 1 meter. This has been anticipated by the IDI mitigation team by recommending a distance of 1 meter and using a barrier between the doctor and patient during the anamnesis. Also, IDI recommends adjusting ventilation and/or airflow to keep doctors safe. However, the reality in the field is that there are still many doctors and/or health centers that have or cannot implement distance, barriers, and ventilation. [9]

### The use of WHO risk assessment

As WHO guidance to assess the risk, it is important to get an early assessment about risk of COVID-19 exposure so that we can intervention how to deal with the result. Assessment risk using WHO questionaiere also done by Rashad in Egypt [10]. In Egypt the result was, there are three clusters of high risk health care workers: 1st group who didn’t wear PPE with infected cases (20%), 2nd group of HCWs who used a PPE but not with all cases or contact with the environment of patients, (20 to 35 %) and 3rd group (34%), of high risk who exposed to accident biological material during interaction with a COVID-19 patient. WHO surveillance raises the staff’s attention to PPE management and recommendation.

### Personal Protection Equipment in Early Pandemic in Indonesia

The situation of the scarcity of PPE in the early pandemic in Indonesia. With the high demand for PPE and the production and distribution at a low level, many physicians are not able to fulfill their PPE needs. Besides that when the scarcity of PPE begins to be felt, as economic principles with high demand, the prices rise, then each doctor tries to fulfill his PPE. Besides the lack of quantity and/or quality of the PPE, many physicians lack the awareness to take precautions. This also fell In China, where at the beginning of the outbreak, there was a general lack of awareness among HCWs to take precautions, and inadequate training among HCWs was noted, with staff incorrectly wearing personal protective equipment (PPE). [11]

In this survey, it was found that only disposable gloves were used more, although the process of changing PPE according to the protocol was carried out by three-quarters of doctors working at the GPHC. This raises the question, is this PPE available at the GPHC? Reflecting on the scarcity of PPE, especially N-95 masks, how do physicians protect themselves. Another study mentions that Indonesian health workers filling their PPE in their own, using their own money (source). Thats why the provision of incentives is expected to meet the needs of PPE and to maintain health. [12]

### Behavior

For behavior, it can be said that almost all respondents have applied hand hygiene and decontamination by breeding. For the behavior of washing hands, Fusaroli et all said that the female gender is more likely to wash their hands properly because of household tasks that make them have to wash their hands. [13]

For surface decontamination, in a study conducted by Samara, it is said that disinfectants are double-edged, if used excessively they will affect the user. [14] Indonesia applies mass spray. It can be concluded that the implementation of disinfection in public areas has the potential to cause health risks [15]. Indonesia has even implemented the manufacture of disinfectant booths to prevent the Covid-19 virus. [16]

### Clasification of Risk

According to the classification that accompanies this questionnaire, it can be seen that 99% of doctors in Indonesia are categorized as high risk. This is also what causes the high mortality rate of doctors. It was far more form the the study the been held by Ali et all. They find evidence suggested that HCWs are being increasingly infected with the novel infection ranging from 15% to 18% and in some cases up to 20% of the infected population. [17]. Furthermore, Ali et all describes the major factors for infection among HCWs include lack of understanding of the disease, inadequate use and availability of Personal Protective Equipment (PPE), uncertain diagnostic criteria, unavailability of diagnostic tests and psychological stress.

## CONCLUSION

First year of pandemic in Indonesia makes the risk for the exposures COVID-19 rises among the physicians. Almost all of the Indonesian Government-owned Public Health Center physicians categorized as high risk. The scarce of personal protection equipment make the phisicians have to strugle to fulfill their PPE needs, more PPE should be produced or imported. With lack of understanding of the disease (described as still doing aerosol-generating procedures and got biological accident), make the training of physicians to identify suspicious cases and to use PPE properly is urgently needed. And the last one is incentives for the Health Workers are needed so they can make complement for the PPE that already provide from health care where they work.

## Data Availability

The datasets used and/or analyzed during the current study are protected by the Ministry of Health of Indonesia and are unsuitable for public sharing

## Author contributions

AS and HH is the main contributor to this manuscript. Study conceptualization and Methodology: AS and HH. Formal analysis: AS. Project administration: AS, TP. Data collection: AS, TP. Funding: AS, TP. Writing (original draft): AS. Writing (review and editing): all authors.

## Ethics statement

The protocol of the study was approved by the Health Research Ethics Committee, National Institute of Health Research and Development, Ministry of Health, Indonesia (LB. 02.01/2/KE.665/2020).

## Data availability statement

The datasets used and/or analyzed during the current study are protected by the Ministry of Health of Indonesia and are unsuitable for public sharing. Interested parties can apply for the data by contacting the data center of the Ministry of Health of Indonesia. All data generated or analyzed during this study are included in this published article.

## Acknowledgment

We would like to thank M. Adib, MD, and the team at the Indonesian Medical Board.

